# Association between Systemic Lupus Erythematosus and Coronary Heart Disease: a retrospective case-control analysis and Mendelian Randomization Study

**DOI:** 10.1101/2021.11.23.21266652

**Authors:** Jinyun Chen, Junmei Tian, Wen Wang, Shiliang Zhou, Lu Zhang, Wanlan Jiang, Mingyuan Cai, Peirong Zhang, Ting Xu, Min Wu

## Abstract

**Objectives:** To appraise the causal effect of systemic lupus erythematosus (SLE) for risk of Coronary heart disease (CHD).

**Methods:** We selected single nucleotide polymorphisms (SNPs) associated with SLE as instrumental variables (IVs) from three independent genome-wide association studies (GWAS), the three largest to date for SLE of European ancestry. Then we conducted two-sample Mendelian randomization (2SMR) analyses to estimate the effects of IVs on the odds of CHD and traditional coronary risk factors (including high LDL cholesterol levels, low HDL cholesterol levels, Apolipoprotein A-I, Apolipoprotein B, diabetes mellitus, and hypertension). Additionally, we searched for common risk loci between SLE and premature coronary atherosclerosis. Furthermore, we retrospectively reviewed the lipid profile of treatment-naïve SLE patients and age-matched healthy controls.

**Results:** Genetically predicted SLE did not increase the odds of CHD. Nevertheless, we found mild causal relationships between SLE and decreased HDL cholesterol levels, and between SLE and decreased apolipoprotein A-I. There was one common risk locus (rs597808) between SLE and premature coronary atherosclerosis at a genome-wide significance level (*P*<5 × 10^−8^). Retrospective analysis showed decreased HDL-cholesterol (0.98±0.516mmol/L vs. 1.46±0.307mmol/L in female, 0.76±0.199mmol/L vs. 1.19±0.257mmol/L in male; both *P*<0.001) and apolipoprotein A-I (1.06±0.314g/L vs. 1.37±0.205g/L in female, 0.87±0.174g/L vs. 1.24±0.200g/L in male; both *P*<0.001) in naïve SLE patients.

**Conclusion:** SLE may accelerate coronary atherosclerosis in young patients by reducing HDL cholesterol and apolipoprotein A-I intrinsically, but it seems not to play a predominant role in CHD development in old patients.

## Introduction

Systemic lupus erythematosus (SLE) is a chronic, multisystem, autoimmune disease with high heritability (λs=29)[1]. Though long-term survival has increased with improved treatments, patients with SLE seem to have substantially increased risk for cardiovascular diseases, including coronary heart disease (CHD), which has been the leading cause of hospitalization and in-hospital mortality in patients with SLE [2–6]. Moreover, the risk of myocardial infarction in SLE patients was higher among Whites compared to Hispanics and Asians [7].

Traditional coronary risk factors such as age, high LDL cholesterol levels, low HDL cholesterol levels, cigarette smoking, diabetes mellitus, and hypertension may partially explain the increase in CHD [8]. Nevertheless, evidence demonstrating that SLE is an independent risk factor for CHD is growing [9–11]. However, most of the previous studies were retrospective observational researches based on small sample sizes, which would be strongly influenced by confounding factors, such as steroid usage. Unconfounded estimates of the causal relationship between SLE and CHD are therefore needed to understand the increased CHD risk in SLE patients.

As we know, SLE is a somewhat hereditary disorder [1, 12], and GWAS has found several risk loci (including SNPs) in SLE. Given that genetic variants are randomly assigned during meiosis according to Mendel’s second law, Mendelian randomization (MR) that uses genetic variants as proxies for exposures to determine the causal relationship between an exposure and an outcome has been developed. We could think of it as a “natural” randomized controlled trial (RCT), which should reduce confounding. Our study aimed to assess the causal relationship between SLE and CHD via MR analysis. In secondary analysis, we also assessed the causal relationship between SLE and traditional coronary risk factors. As patients with SLE were found to have premature coronary atherosclerosis, we also searched for common risk loci between SLE and premature coronary atherosclerosis. To confirm the results with real-world data, we retrospectively reviewed the lipid profile and the prevalence of CHD, diabetes mellitus and hypertension in treatment-naïve SLE patients.

## Methods

### Study populations

The MR analysis included three GWAS on SLE with summary-level data (Table 1). The study also included large sizes of participants enrolled in CARDIoGRAMplusC4D Consortium or CARDIoGRAM Consortium, two meta-analyses of GWAS on coronary heart disease for whom summary-level data were available (mean age>50 years old) [13, 14]. GWAS on hypertension, diabetes mellitus, LDL cholesterol levels, HDL cholesterol levels, apolipoprotein A-I, and apolipoprotein B were included as well. All of the participants were of European ancestry (except for CARDIoGRAMplusC4D Consortium, which was consisted of 92.85% European and 7.15% South Asian ancestry) and provided written informed consent in each of the contributing studies. We acquired summary data for all SNPs by our search from NHGRI-EBI GWAS catalog[15] or MR Base database [16]. The risk loci associated with premature coronary atherosclerosis were acquired from genome-wide scale time-to-event data analysis of UK Biobank data (282,871 British European ancestry samples) [17].

**Table 1.** Description of Contributing Studies.

| **Contributing Studies**<br>**(Consortium or Authors)** | **Sample Size**<br>**(Cases/Controls)** | **Ancestry** |
| --- | --- | --- |
| **GWAS for SLE** |  |  |
| Bentham, J. et al. | 23,210(7,219/15,991) | European |
| Langefeld, C.D. et al. | 18,264(6,748/11,516) | European |
| Gateva, V. et al. | 15,461(3,273/12,188) | European |
| **GWAS for CHD** |  |  |
| CARDIoGRAMplusC4D | 194,427(63,746/130,681) | 92.85% European, 7.15% South Asian |
| CARDIoGRAM | 86,995(22,233/64,762) | European |
| **GWAS for risk factors for CHD** |  |  |
| Hypertension (MRC-IEU) | 463,010(54,358/408,652) | European |
| Diabetes (MRC-IEU) | 461,578(22,340/439,238) | European |
| HDL-cholesterol (Kettunen, J. et al.) | 21,555 | European |
| LDL-cholesterol (Kettunen, J. et al.) | 21,559 | European |
| Apolipoprotein A-I (Kettunen, J. et al.) | 20,687 | European |
| Apolipoprotein B (Kettunen, J. et al.) | 20,690 | European |

The real-world data were derived from treatment-naïve SLE patients who were still followed-up in our hospital in the last two years and age-matched controls who had a health check in the same period. We retrospectively reviewed the lipid profile of SLE patients (n=218) and the healthy controls (n=95,132). The prevalence of CHD, diabetes mellitus and hypertension was evaluated in those SLE patients as well (Table 3).

### Patient and Public Involvement

Patients or the public were not involved in the design, or conduct, or reporting, or dissemination plans of our research.

### Instrumental variables

We searched the GWAS catalog on March 8, 2020, to identify SNPs associated with SLE (using a *P*-value threshold of 5 × 10^−8^), and exclude those that were in significant linkage disequilibrium (r^2^_>_0.3). If a particular SNP is not present in the outcome dataset, an LD proxy would be used instead (within 250 kb and with a minimum r^2^= 0.8). Using the remaining SNPs or LD proxies as independent instrumental variables (IVs), we applied 2-sample Mendelian randomization to estimate the effect of SLE on CHD or traditional coronary risk factors.

### Statistical analysis

Inference of causality in the estimated etiological associations between SLE and CHD or traditional coronary risk factors depends on the satisfaction of Mendelian randomization assumptions[18]: (1) the selected SNPs are associated with SLE; (2) the selected SNPs are not associated with confounders; and (3) the selected SNPs are associated with CHD or traditional coronary risk factors exclusively through their effect on SLE. If the above assumptions are satisfied, the selected SNPs are valid IVs, and their association with CHD or traditional coronary risk factors can be interpreted as a causal effect of SLE.

The association between genetically predicted SLE and CHD or traditional coronary risk factors attributable to each SNP was accessed with the Wald method, which computes the ratio between the SNP-SLE and SNP-CHD or SNP-traditional coronary risk factors estimates. The ratio estimates for individual SNPs were combined by using the inverse-variance weighted (IVW) meta-analysis method (random-effects). We additionally examined the violation of the MR assumptions through sensitivity analyses, based on the weighted median, MR-Egger, and weighted mode approaches. CARDIoGRAMplusC4D Consortium was set as the outcome in MR analysis unless there was directional horizontal pleiotropy, when CARDIoGRAM Consortium would replace it. Cochran’s Q value was furthermore estimated to measure the heterogeneity in the inverse-variance weighted (IVW) and MR-Egger analysis. If there was significant heterogeneity between SNPs, they would be further analyzed with outlier-corrected MR-PRESSO.

The prevalence of SLE in Caucasians was estimated at 47.5 per 100,000 persons[19]. Effect allele frequencies of SNPs were estimated with SNPsnap[20] using 1000 Genomes data if not available from GWAS. And heritability of liability attributable to each SNP (*h*^2^) was estimated with INDI-V[21].

All the above analyses were performed in R, version 3.5.1. Lipid profile in SLE patients was compared with that in controls via unpaired t-test. *P* values were 2-sided, and evidence of association was declared at *P*<0.05.

## Results

### Genetic loci associated with SLE

The identified risk loci in the three GWAS for SLE explained an estimated 8% -19.3% [22, 23]of the total genetic susceptibility to SLE. We selected SNPs (or LD proxies) from these three GWAS as IVs for SLE risk (Table 1 and Supplementary Table S1). The selected SNPs correspond to independent genomic regions with an odds ratio (OR) from 1.13 to 2.28, and an *h*^2^ from 0.06% to 3.28% for SLE.

### Associations of genetic risk of SLE with coronary heart disease

To estimate the effect of genetically raised SLE risk on CHD risk, we used the IVW MR analysis (random-effects) in which the SLE risk-increasing variants were treated as IVs (Figure 1 and Methods section). A random-effects IVW model yielded a pooled MR estimate of nonsignificant effect, demonstrating that SLE increased the odds of CHD by 1.00 (95% CI, 0.97, 1.04; *P*= 0.91, Bentham, J. et al.), 1.01 (95% CI, 0.98, 1.04; *P*= 0.63, Langefeld, C.D. et al.), and 0.98 (95% CI, 0.94, 1.02; *P*= 0.24, Gateva, V. et al.), respectively (Table 2, Figure 1). Weighted median and weighted mode estimates for the candidate SNPs yielded results similar to those of the random-effects IVW model. We did not find the presence of directional horizontal pleiotropy in any of the three analyses, as indicated by the MR-Egger intercept test (*P*= 0.25, 0.76, and 0.53, respectively). There was no evidence of heterogeneity in the IVW and MR-Egger analysis with IVs from GWAS by Langefeld, C.D. et al. (Q=3.91, *P*=0.56; Q=3.81, *P*=0.43, respectively) and Gateva, V. et al. (Q=5.76, *P*=0.33; Q=5.16, *P=*0.27, respectively). However, with IVs from GWAS by Bentham J et al., there was significant heterogeneity (Q=66.22, *P*<0.001; Q=63.36, *P*<0.001, respectively), which was further analyzed with outlier-corrected MR-PRESSO (Figure 1).

**Figure 1.**
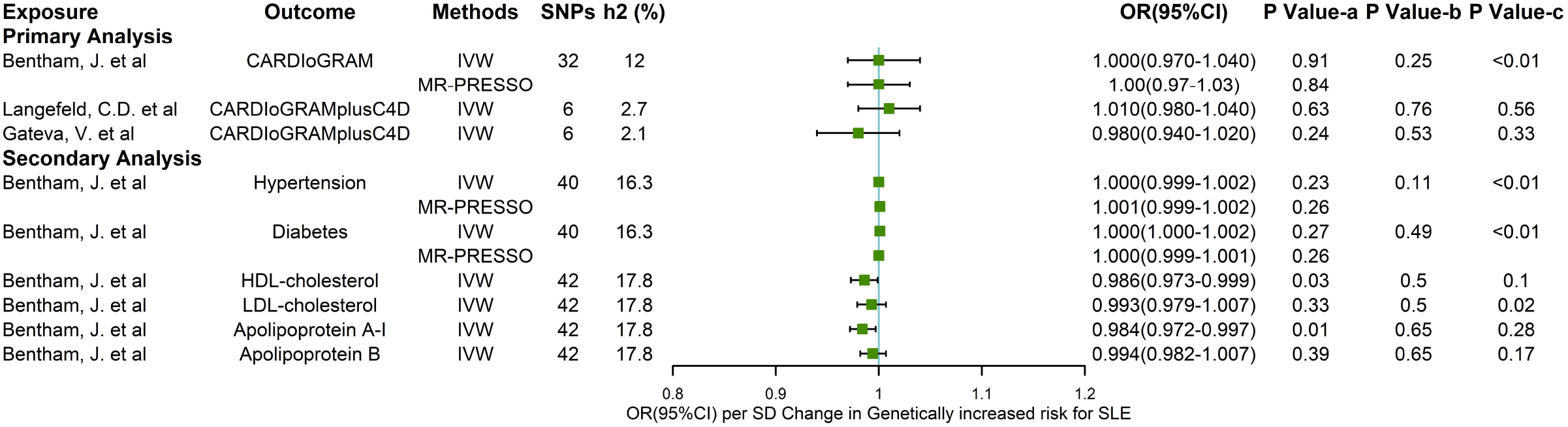
Associations between genetically increased SLE risk and odds of CHD or traditional risk factors for CHD. Mendelian randomization analyses of the causal relationship between SLE and CHD or traditional risk factors for CHD in the inverse-variance weighted model. SNPs, single nucleotide polymorphisms; h2, heritability of liability attributable to this risk variant by the approximate equation; OR, odds ratio. *P* value-a, *P*-value for the association between genetically increased SLE risk and diseases from IVW meta-analysis (random-effects). *P* value-b, *P* value for horizontal Pleiotropy from MR-Egger intercept test. *P* value-c, *P* value for heterogeneity among SNPs within the instrument from Cochran’s Q test.

**Table 2.**
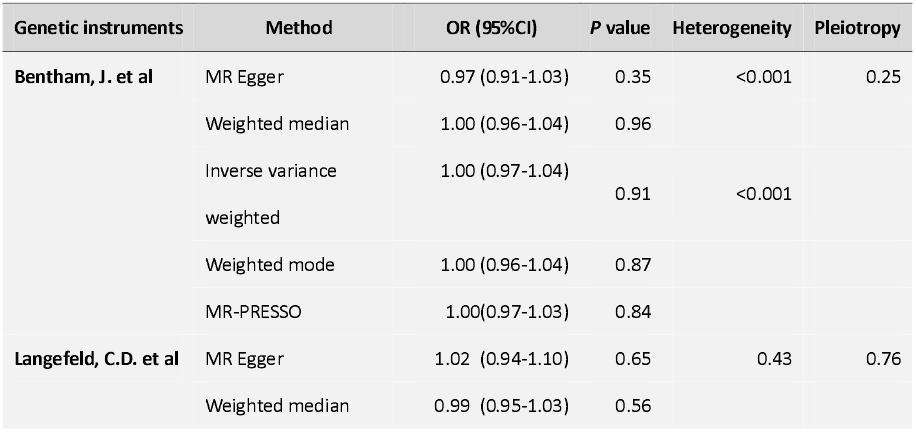
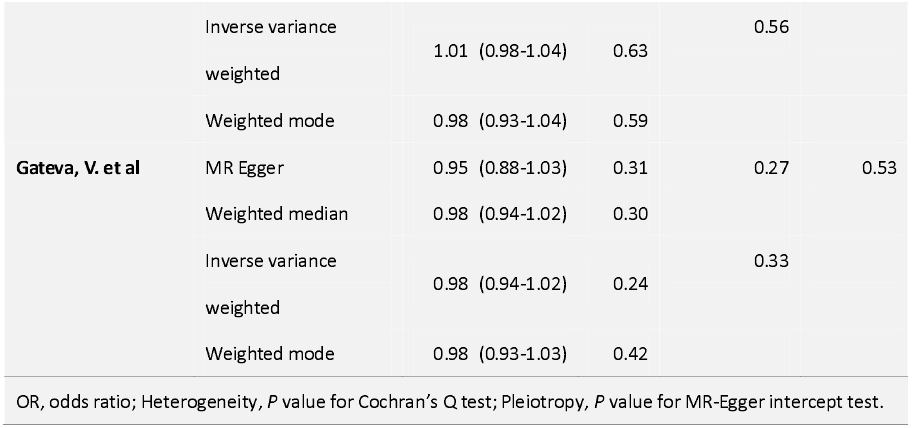
Two-sample MR estimates for the effect of SLE on CHD risk.

### Associations of genetic risk of SLE with traditional coronary risk factors

MR analysis (IVW random-effects) indicated that SLE increased the odds of LDL-cholesterol by 0.99 (95% CI, 0.98, 1.01; *P*= 0.33), HDL-cholesterol by 0.99 (95% CI, 0.97, 1.00; *P*= 0.03), apolipoprotein A-I by 0.98 (95% CI, 0.97, 1.00; *P*= 0.01), apolipoprotein B by 0.99 (95% CI, 0.98, 1.01; *P*= 0.39), diabetes mellitus by 1.001 (95% CI, 1.000, 1.001; *P*= 0.27) and hypertension by 1.001 (95% CI, 1.000, 1.002; *P*= 0.23), respectively, without directional horizontal pleiotropy (Figure 1).

### Association between SLE and premature coronary atherosclerosis

A genome-wide scale time-to-event data analysis of UK Biobank data identified 69 risk loci associated with premature coronary atherosclerosis [17]. There was one common risk locus (rs597808-A) between SLE and premature coronary atherosclerosis at a genome-wide significance level (*P*<5 × 10^−8^), which was also associated with decreased HDL and LDL [24].

### Dyslipidemia in treatment-naïve SLE patients

We reviewed the baseline lipid profile before any treatments in 218 SLE patients (198 female/20 male). An age-matched population (45,613 female/49,519 male) were involved as healthy controls. In both sexes, HDL-cholesterol (0.98±0.516mmol/L vs. 1.46±0.307mmol/L in female, 0.76±0.199mmol/L vs. 1.19±0.257mmol/L in male; both *P*<0.001) and apolipoprotein A-I (1.06±0.314g/L vs. 1.37±0.205g/L in female, 0.87±0.174g/L vs. 1.24±0.200g/L in male; both *P*<0.001) were significantly decreased in naïve SLE patients. Additionally, LDL-cholesterol, apolipoprotein B, and total cholesterol were decreased as well. Triglyceride level was increased in female SLE patients, but not in male patients (Table 3). The prevalence of CHD, diabetes mellitus, and hypertension in SLE patients was 0.9%, 1.4%, and 11.9%, respectively, which were comparable to or lower than that in the general population in China.

**Table 3.**
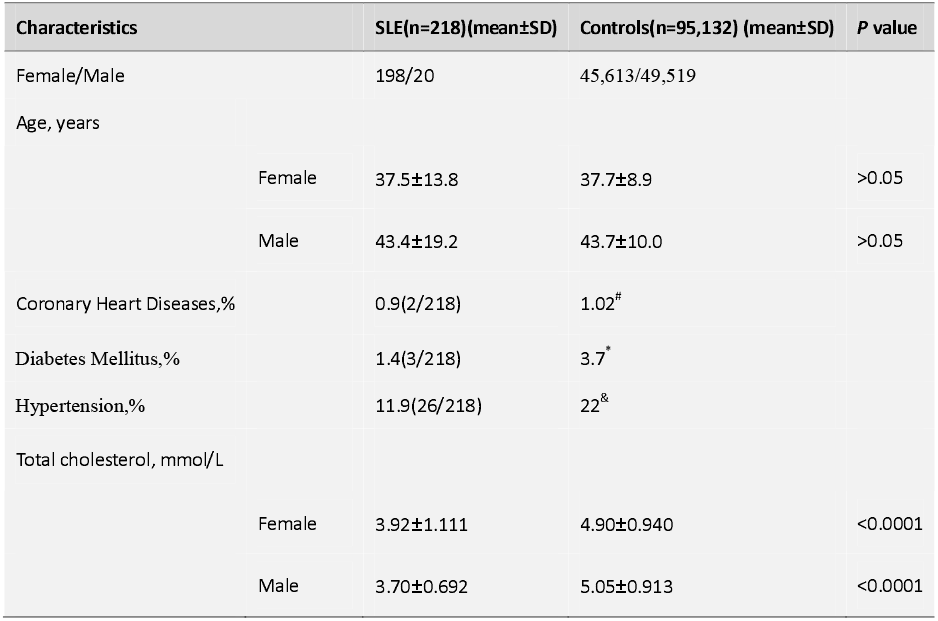
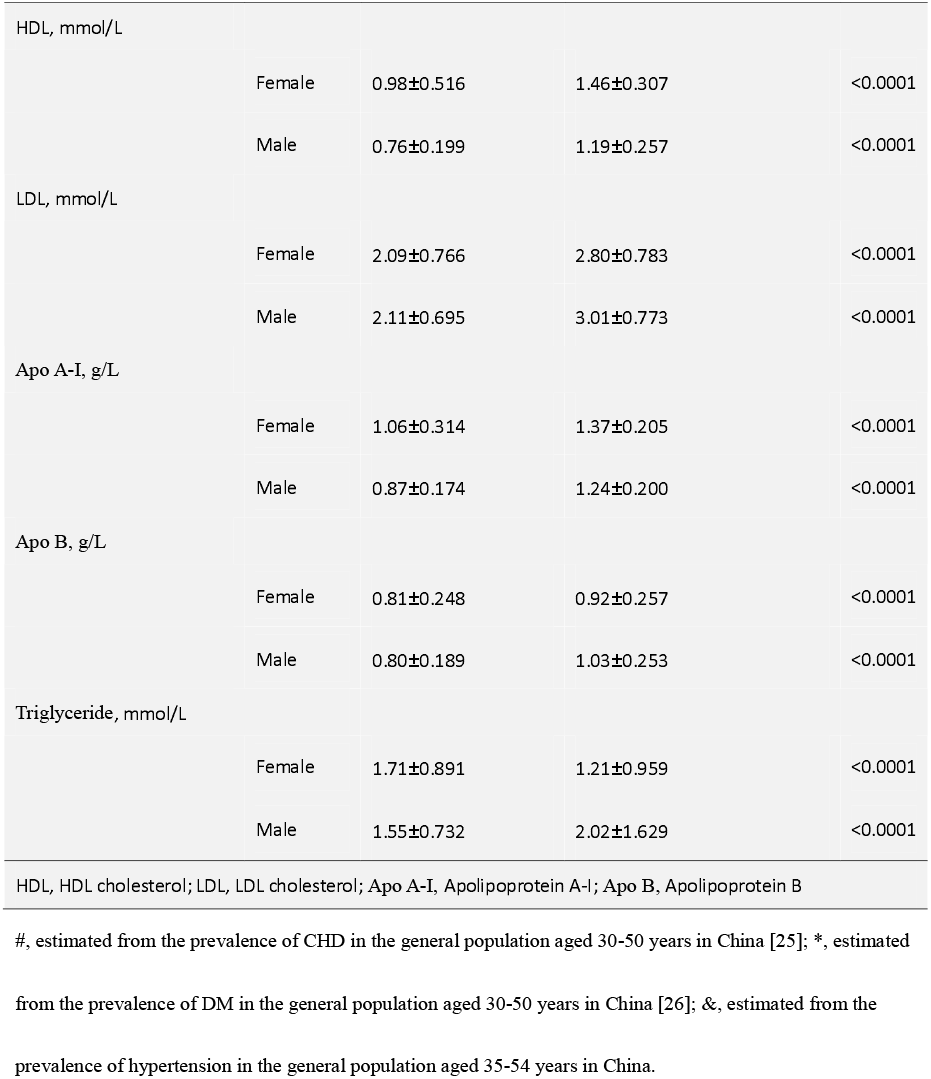
Characteristics and lipid profiles in naïve SLE patients and healthy controls.

## Discussion

Our study did not find a significant causal effect of SLE on CHD, indicating that genetically predicted SLE may not cause coronary heart disease in old patients (>50 years old). Nevertheless, the secondary analysis showed that SLE had a mild decreasing effect on HDL-cholesterol and apolipoprotein A-I, which was consistent with previous observational studies[27] and our real-world data analysis. It may partially explain premature coronary-artery atherosclerosis in young SLE patients[6].

Previous investigations demonstrated an increased risk for CHD in SLE patients, in which both traditional and SLE-specific risk factors are important [8, 11]. However, there are still discrepancies within the literature. Some studies showed similar disease activity in SLE patients with or without coronary-artery calcification, and accelerated plaque progression in SLE regardless of disease activity [6, 10], while others showed SLE disease activity as one of the predictors of CHD [9, 28]. Additionally, previous studies diverged regarding whether atherosclerosis was more common in SLE than in controls, and further subgroup analyses found that lupus nephritis (LN) accelerated atherosclerosis and increased risk for CHD [4, 29].

Because of the existence of several closely associated risk factors, such as disease activity and glucocorticoid use, it is difficult to disentangle their effects [30]. For instance, a recent study compared atherosclerotic comorbidities in corticosteroid-naive and corticosteroid-exposed patients with SLE, with a minimum of 3 years of follow-up[31]. They found no difference in the frequency of coronary artery disease overall or at each of the time points (3, 5, and 8 years, respectively). However, the corticosteroid-exposed patients had higher Systemic Lupus Erythematosus Disease Activity Index 2000 scores on first examination and more immunosuppressive treatment, which were not balanced between the two groups.

Nevertheless, cumulative corticosteroid dose and disease duration were found to be associated with plaque progression in most of the studies [9, 10]. A recent large matched cohort study showed that the risk for CHD was highest during the first year after SLE diagnosis [32].

Moreover, a long-term prospective study indicated that disease-related factors seemed to dominate CHD risk during the first 8 years, while traditional factors such as hypertension and diabetes, partially related to corticosteroid treatment, played an important role later in the disease course [33]. Notably, increased CHD prevalence in SLE was more common in younger patients rather than in old patients (>50 years old). The association between SLE and CHD was attenuated with increasing age [3].

As a complex trait, CHD would be influenced by lots of factors besides genetics, especially in old patients. As we know, CHD commonly occurred in old patients who were more than 50 years old. However, the onset of SLE was mainly in young females around child-birth age. The age gap may dilute the genetic effect of SLE on CHD. So, despite the null result of our primary analysis, we could not draw the same conclusion in young-onset CHD patients. In contrast, we may get a different result if calculating the effect of IVs for SLE against GWAS of CHD in young patients, as demonstrated by the common risk locus (rs597808-A) between SLE and premature coronary atherosclerosis in the time-to-event data analysis of UK Biobank data. Further analyses are needed to take into account the age gap between the onset of SLE and CHD in the general population.

Besides HDL-cholesterol and apolipoprotein A-I, LDL-cholesterol, apolipoprotein B, and total cholesterol were decreased in naïve SLE patients as well. Rs597808 may partly account for the dyslipidemia in SLE [24, 34]. However, it still needs further investigation to elucidate whether and how the dyslipidemia participated in the premature coronary atherosclerosis in SLE.

The limitations of this study include five parts. First, data of associations of SNP-exposure and SNP-outcome were derived from 2 different populations, which would increase heterogeneity, as we saw in the SNPs from GWAS by Bentham, J. et al. Second, we included SNPs only those with a genome-wide significance level (*P*<5 × 10^−8^), meaning those genuinely associated variants that did not reach the stringent p-value threshold were ignored. Recently, e.g., a comparative study found that an IL19 risk allele, rs17581834 (T), was associated with stroke/myocardial infarction in SLE (OR 2.3 (1.5 to 3.4), *P*=8.5×10^−5^). However, we neither found it in the three GWAS on SLE nor in the CARDIoGRAMplusC4D Consortium dataset due to *P*-value. Third, though three GWAS on SLE were all of European ancestry, CARDIoGRAMplusC4D Consortium was of mixed ancestry (92.85% European and 7.15% South Asian). However, the directional horizontal pleiotropic effect was not observed in the MR-Egger regression in this study. Fourth, we may have underestimated the risk of CHD in SLE, since there may be an increased risk for early death from CHD in SLE. Last, the real-world data were from the Chinese population rather than the European population. Despite the difference, the majority of genetic risk polymorphisms for SLE are present in both populations[35], supporting the effectiveness of the analysis.

Though most of the selected SNPs were each estimated to account for less than 1% of the genetic variance, multi-SNPs in combination, i.e., IVs, could explain up to 12% of estimated heritability of liability to SLE in the primary analysis (16.3-17.8% in the secondary analysis). Additionally, we conducted 2-sample MR analyses by using genetic instruments from three large independent GWAS separately in our study, which would strengthen the power of our conclusion.

**Key messages**

**What is already known about this subject?**

Patients with systemic lupus erythematosus (SLE) has increased risk for coronary heart disease (CHD), which has been the leading cause of hospitalization and in-hospital mortality in patients with SLE. But whether there is a causal relationship between SLE and CHD is uncertain. Genetic variants related to SLE can be used as proxies for SLE to help judge causality (“Mendelian randomisation analyses”). Previous studies have been insufficiently powerful and detailed to evaluate the possibility of any causal role for SLE in CHD.

**What does this study add?**

SLE may accelerate coronary atherosclerosis in young patients by reducing HDL cholesterol and apolipoprotein A-I intrinsically, but it seems not to play a predominant role in CHD development in old patients.

**How might this impact on clinical practice or future developments?**

We should caution the dyslipidemia and premature coronary atherosclerosis in young SLE patients. Treatments for SLE, such as corticosteroid use, should be considered more carefully to decrease CHD risk.

## Data Availability

The data underlying this article are available in the NHGRI-EBI GWAS catalog, at https://www.ebi.ac.uk/gwas/, and MR Base database, at http://app.mrbase.org. The cumulative risk curves of the risk loci for coronary artery atherosclerosis are available at ftp://share.sph.umich.edu/UKBB_SPACox_HRC/Survplot.

## Acknowledgements

We wish to thank the CARDIOGRAMplusC4D, CARDIOGRAM, MRC-IEU, GWAS by Bentham, J. et al., Langefeld, C.D. et al., Gateva, V. et al., Kettunen, J. et al., and Wenjian, B. et al. for access to their data.

## Funding

Supported by grants from the Science and Technology Project of Changzhou Health Committee for Young Talents (QN201805).

## Contributors

The corresponding author attests that all listed authors meet authorship criteria and that no others meeting the criteria have been omitted.

## Competing interests

None declared.

## Ethical approval

The study was conducted in accordance with the Declaration of Helsinki (as revised in 2013). This study was approved by the Ethics Committee of Soochow University and individual consent for this retrospective analysis was waived.

## Notes

### Competing Interest Statement

The authors have declared no competing interest.

### Author Declarations

This study was approved by the Ethics Committee of Soochow University and individual consent for this retrospective analysis was waived.

