## supplemental Table 1 for "Association between Systemic Lupus Erythematosus and Coronary Heart Disease: a retrospective case-control analysis and Mendelian Randomization Study"

**Dyslipidemia and Coronary Heart Disease in Patients with Systemic Lupus Erythematosus: a real-world and Mendelian Randomization Study**

**Jinyun Chen^1^, Junmei Tian^1^, Wen Wang^1^, Shiliang Zhou^1^, Lu Zhang^1^, Wanlan Jiang^1^, Mingyuan Cai^1^, Peirong Zhang^1^, *Ting Xu^1^ & *Min Wu^1^**

**Jinyun Chen,** M.D. & Ph.D.*,* **^1^**Department of Rheumatology and Immunology, Third Affiliated Hospital of Soochow University, Changzhou 213000, China.;

**Junmei Tian,** M.D.*,* Department of Rheumatology and Immunology, Third Affiliated Hospital of Soochow University, Changzhou 213000, China.;

**Wen Wang,** M.D.*,* Department of Rheumatology and Immunology, Third Affiliated Hospital of Soochow University, Changzhou 213000, China.;

**Shiliang Zhou,** M.D.*,* Department of Rheumatology and Immunology, Third Affiliated Hospital of Soochow University, Changzhou 213000, China.;

**Lu Zhang,** M.D.*,* Department of Rheumatology and Immunology, Third Affiliated Hospital of Soochow University, Changzhou 213000, China.;

**Wanlan Jiang,** M.D.*,* Department of Rheumatology and Immunology, Third Affiliated Hospital of Soochow University, Changzhou 213000, China.;

**Mingyuan Cai,** M.D.*,* Department of Rheumatology and Immunology, Third Affiliated Hospital of Soochow University, Changzhou 213000, China.;

**Peirong Zhang,** M.D.*,* Department of Rheumatology and Immunology, Third Affiliated Hospital of Soochow University, Changzhou 213000, China.;

**Ting Xu,** M.D. & Ph.D.*,* Department of Rheumatology and Immunology, Third Affiliated Hospital of Soochow University, Changzhou 213000, China.;

**Min Wu,** M.D. & Ph.D.*,* Department of Rheumatology and Immunology, Third Affiliated Hospital of Soochow University, Changzhou 213000, China.;

* Correspondence and requests for materials should be addressed to T.X. or M.W..

**Funding:** Supported by grants from the Science and Technology Project of Changzhou Health Committee for Young Talents (QN201805, QN201910).

**Supplementary Table S1 | Genome wide significant single nucleotide polymorphisms (SNPs) for SLE**

| **SNP** | **Location** | **Mapped gene** | **EA** | **EAF** | **OR** | **95%CI** | **h2 (%)** | **P Value** |
| --- | --- | --- | --- | --- | --- | --- | --- | --- |
| **Bentham, J. et al** | |  |  |  |  |  |  |  |
| rs10028805 | 4:101816093 | BANK1 | G | 0.63 | 1.2 | [1.15–1.25] | 0.21 | 4E-17 |
| rs10036748 | 5:151078585 | TNIP1 | T | 0.26 | 1.38 | [1.32–1.45] | 0.62 | 1E-45 |
| rs10488631 | 7:128954129 | AC025594.2, TNPO3 | C | 0.10 | 1.92 | [1.81–2.03] | 1.72 | 9E-110 |
| rs1059312 | 12:128794319 | SLC15A4 | C | 0.42 | 1.17 | [1.12–1.21] | 0.16 | 1E-13 |
| rs10774625 | 12:111472415 | ATXN2 | A | 0.48 | 1.13 | [1.08–1.18] | 0.10 | 4E-09 |
| rs11644034 | 16:85939006 | AC092723.5 | G | 0.81 | 1.25 | [1.19–1.32] | 0.22 | 1E-17 |
| rs11889341 | 2:191079016 | STAT4 | T | 0.23 | 1.73 | (1.65–1.81] | 2.11 | 6E-122 |
| rs1270942 | 6:31951083 | CFB, AL645922.1 | C | 0.10 | 2.28 | [2.15–2.42] | 3.17 | 2E-165 |
| rs12802200 | 11:566936 | MIR210HG | C | 0.81 | 1.23 | [1.15–1.31] | 0.19 | 9E-10 |
| rs1734787 | X:154059995 | MECP2 | C | 0.18 | 1.31 | [1.22–1.40] | 0.32 | 2E-15 |
| rs17849501 | 1:183573188 | SMG7, NCF2 | T | 0.06 | 2.1 | [1.95–2.26] | 1.53 | 3E-88 |
| rs1801274 | 1:161509955 | FCGR2A | C | 0.49 | 1.16 | [1.11–1.21] | 0.15 | 1E-12 |
| rs2111485 | 2:162254026 | FAP, IFIH1 | G | 0.40 | 1.15 | [1.11–1.20] | 0.12 | 1E-11 |
| rs2286672 | 17:4809322 | PLD2 | T | 0.09 | 1.25 | [1.16–1.35] | 0.11 | 3E-09 |
| rs2289583 | 15:75018695 | SCAMP5 | A | 0.31 | 1.19 | [1.14–1.24] | 0.18 | 6E-15 |
| rs2304256 | 19:10364976 | TYK2 | C | 0.74 | 1.24 | [1.17–1.31] | 0.25 | 4E-13 |
| rs2431697 | 5:160452971 | MIR3142HG | T | 0.57 | 1.26 | [1.21–1.31] | 0.38 | 8E-28 |
| rs2476601 | 1:113834946 | AL137856.1, PTPN22 | A | 0.09 | 1.43 | [1.34–1.53] | 0.36 | 1E-28 |
| rs2663052 | 10:48861350 | WDFY4 | C | 0.51 | 1.16 | [1.10–1.22] | 0.15 | 5E-09 |
| rs2732549 | 11:35066852 | AL356215.1, PDHX | T | 0.56 | 1.24 | [1.19–1.29] | 0.32 | 1E-23 |
| rs2736340 | 8:11486464 | BLK, AF131216.5 | T | 0.24 | 1.29 | [1.22–1.37] | 0.35 | 6E-20 |
| rs2941509 | 17:39764941 | IKZF3 | A | 0.03 | 1.35 | [1.22–1.49] | 0.08 | 8E-09 |
| rs3024505 | 1:206766559 | IL10, AL591846.1 | T | 0.17 | 1.17 | [1.11–1.24] | 0.09 | 5E-09 |
| rs34572943 | 16:31261032 | ITGAM | A | 0.13 | 1.71 | [1.61–1.81] | 1.27 | 3E-76 |
| rs3768792 | 2:213006985 | AC093865.1, IKZF2 | C | 0.14 | 1.24 | [1.17–1.31] | 0.16 | 1E-13 |
| rs3794060 | 11:71476633 | NADSYN1 | C | 0.30 | 1.23 | [1.18–1.29] | 0.25 | 1E-20 |
| rs4902562 | 14:68264741 | RAD51B | A | 0.39 | 1.14 | [1.09–1.19] | 0.11 | 6E-10 |
| rs4917014 | 7:50266267 | AC020743.3, AC020743.2 | T | 0.65 | 1.18 | [1.13–1.24] | 0.17 | 6E-14 |
| rs4948496 | 10:62045858 | ARID5B | C | 0.50 | 1.14 | [1.10–1.19] | 0.11 | 1E-10 |
| rs564799 | 3:160011200 | IL12A-AS1 | C | 0.65 | 1.14 | [1.09–1.18] | 0.10 | 2E-09 |
| rs6568431 | 6:106140931 | ATG5 | A | 0.39 | 1.21 | [1.15–1.27] | 0.24 | 5E-14 |
| rs6932056 | 6:137921300 | LINC02528, AL591468.1 | C | 0.02 | 1.83 | [1.65–2.02] | 0.31 | 2E-31 |
| rs704840 | 1:173257056 | TNFSF4, AL645568.3 | G | 0.28 | 1.22 | [1.17–1.27] | 0.22 | 3E-19 |
| rs7444 | 22:21622645 | UBE2L3 | C | 0.20 | 1.27 | [1.21–1.33] | 0.26 | 2E-22 |
| rs7726414 | 5:134096143 | TCF7, AC008608.1 | T | 0.07 | 1.45 | [1.32–1.58] | 0.30 | 4E-16 |
| rs7941765 | 11:128629105 | AP001122.1 | C | 0.50 | 1.14 | [1.10–1.19] | 0.11 | 1E-10 |
| rs849142 | 7:28146272 | JAZF1 | A | 0.51 | 1.14 | [1.10–1.19] | 0.11 | 9E-11 |
| rs887369 | X:30559729 | CXorf21 | C | 0.79 | 1.15 | [1.10–1.21] | 0.09 | 5E-10 |
| rs9311676 | 3:58484624 | AC116036.2, PDHB | C | 0.59 | 1.17 | [1.13–1.22] | 0.16 | 3E-14 |
| rs9462027 | 6:34829464 | UHRF1BP1 | A | 0.29 | 1.14 | [1.09–1.19] | 0.09 | 8E-09 |
| rs9652601 | 16:11080508 | CLEC16A | G | 0.67 | 1.21 | [1.15–1.26] | 0.22 | 7E-17 |
| rs9782955 | 1:235876577 | LYST | C | 0.77 | 1.16 | [1.11–1.22] | 0.10 | 1E-09 |
| **Langefeld, C.D. et al** | | |  |  |  |  |  |  |
| rs1143679 | 16:31265490 | ITGAM | A | 0.11 | 1.72 | [1.61-1.83] | 1.13 | 3E-62 |
| rs12129787 | 1:161522797 | AL590385.2, FCGR2A | A | 0.35 | 1.22 | [1.14-1.29] | 0.25 | 1E-09 |
| rs35789010 | 6:25513951 | CARMIL1 | A | 0.07 | 1.46 | [1.35-1.59] | 0.32 | 5E-19 |
| rs36014129 | 6:25884291 | H2BP5, H2AC3P | A | 0.08 | 1.5 | [1.39-1.62] | 0.41 | 1E-24 |
| rs6679677 | 1:113761186 | RSBN1, PHTF1 | A | 0.10 | 1.41 | [1.32-1.51] | 0.35 | 2E-23 |
| rs11059927 | 12:128809788 | SLC15A4 | C | 0.10 | 1.21 | [1.13-1.30] | 0.09 | 2E-08 |
| rs1131265 | 3:119503609 | TIMMDC1 | C | 0.81 | 1.23 | [1.15-1.32] | 0.19 | 1E-09 |
| rs180977001 | 3:58332737 | PXK, AC098479.1 | C | 0.07 | 1.27 | [1.17-1.39] | 0.10 | 2E-08 |
| rs2327832 | 6:137651931 | AL356234.2, BTF3L4P3 | C | 0.21 | 1.22 | [1.15-1.28] | 0.18 | 2E-13 |
| rs2980512 | 8:8283379 | PRAG1, FAM86B3P | C | 0.47 | 1.15 | [1.10-1.20] | 0.13 | 4E-10 |
| rs7582694 | 2:191105394 | STAT4 | C | 0.23 | 1.56 | [1.48-1.64] | 1.25 | 4E-69 |
| rs7819602 | 8:10869332 | XKR6, AC011008.2 | C | 0.39 | 1.15 | [1.10-1.20] | 0.12 | 1E-09 |
| rs12575600 | 11:128454974 | LINC02098, ETS1 | G | 0.10 | 1.24 | [1.16-1.33] | 0.12 | 6E-10 |
| rs131658 | 22:21563337 | UBE2L3 | G | 0.20 | 1.25 | [1.19-1.32] | 0.23 | 1E-16 |
| rs3122605 | 1:206781696 | IL19 | G | 0.14 | 1.23 | [1.16-1.30] | 0.15 | 1E-11 |
| rs10245867 | 7:28102567 | JAZF1 | T | 0.33 | 1.14 | [1.09-1.19] | 0.10 | 4E-08 |
| rs12706861 | 7:128976528 | TNPO3 | T | 0.12 | 1.76 | [1.65-1.87] | 1.36 | 4E-71 |
| rs2299864 | 6:106220119 | ATG5 | T | 0.20 | 1.24 | [1.17-1.30] | 0.21 | 6E-15 |
| rs2736336 | 8:11484361 | AF131216.5, BLK | T | 0.25 | 1.34 | [1.28-1.41] | 0.49 | 6E-32 |
| rs4690229 | 4:976936 | DGKQ | T | 0.46 | 1.13 | [1.09-1.19] | 0.10 | 2E-08 |
| rs77000060 | 6:137916852 | LINC02528, AL591468.1 | T | 0.03 | 1.89 | [1.69-2.11] | 0.52 | 2E-29 |
| rs9462027 | 6:34829464 | UHRF1BP1 | T | 0.27 | 1.15 | [1.09-1.20] | 0.10 | 2E-08 |
| **Gateva, V. et al** | |  |  |  |  |  |  |  |
| rs11755393 | 6:34856859 | UHRF1BP1 | G | 0.35 | 1.17 | [1.10–1.24] | 0.15 | 2E-08 |
| rs11860650 | 16:31315385 | ITGAM | T | 0.13 | 1.43 | [1.32–1.54] | 0.48 | 2E-20 |
| rs2070197 | 7:128948946 | IRF5 | C | 0.11 | 1.88 | [1.78–1.95] | 1.71 | 6E-24 |
| rs2205960 | 1:173222336 | AL645568.3, TNFSF4 | T | 0.23 | 1.22 | [1.15–1.30] | 0.20 | 6E-09 |
| rs2476601 | 1:113834946 | AL137856.1, PTPN22 | A | 0.1 | 1.35 | [1.24–1.47] | 0.25 | 3E-12 |
| rs2736340 | 8:11486464 | BLK, AF131216.5 | T | 0.25 | 1.35 | [1.27–1.43] | 0.52 | 8E-17 |
| rs3024505 | 1:206766559 | IL10, AL591846.1 | A | 0.16 | 1.19 | [1.11–1.28] | 0.11 | 4E-08 |
| rs3135394 | 6:32440720 | HLA-DRA | G | 0.1 | 1.98 | [1.84–2.14] | 1.94 | 2E-60 |
| rs4963128 | 11:589564 | PHRF1 | C | 0.67 | 1.2 | [1.13–1.27] | 0.20 | 5E-09 |
| rs5029937 | 6:137874014 | TNFAIP3 | T | 0.03 | 1.71 | [1.51–1.95] | 0.33 | 5E-13 |
| rs6568431 | 6:106140931 | ATG5 | A | 0.38 | 1.2 | [1.14–1.27] | 0.21 | 7E-10 |
| rs7574865 | 2:191099907 | STAT4 | T | 0.23 | 1.57 | [1.49–1.69] | 1.30 | 1E-41 |
| rs7708392 | 5:151077924 | TNIP1 | C | 0.24 | 1.27 | [1.10–1.35] | 0.30 | 4E-13 |
| rs849142 | 7:28146272 | JAZF1 | T | 0.49 | 1.19 | [1.13–1.26] | 0.21 | 2E-09 |
| EA, effect allele; EAF, effect allele frequency; OR, odds ratio; h2, heritability of liability attributable to this risk variant by the approximate equation. | | | | | | | | |
